# Cannabis use and the risk of primary open-angle glaucoma: a Mendelian randomization study

**DOI:** 10.1101/2022.12.15.22283517

**Authors:** Andreas Katsimpris, Sebastian-Edgar Baumeister, Hansjörg Baurecht, Andrew Tatham, Michael Nolde

**Affiliations:** Department of Ophthalmology, Aberdeen Royal Infirmary, Aberdeen, AB25 2ZN, UK; Institute of Health Services Research in Dentistry, University of Münster, Münster, Germany; Department of Epidemiology and Preventive Medicine, University of Regensburg, Regensburg, Germany; Princess Alexandra Eye Pavilion, Edinburgh, EH16 5XR, UK

## Abstract

**Background:** Several observational studies have investigated the association between cannabis use and intraocular pressure, but its association with primary open-angle glaucoma (POAG) remains unclear. In this study, we leveraged human genetic data to assess through Mendelian randomization (MR) whether cannabis use affects POAG.

**Methods:** We used five single-nucleotide polymorphisms (SNPs) associated with lifetime cannabis use (P-value < 5×10^−8^) from a genome-wide association study (GWAS) (N = 184,765) by the International Cannabis Consortium, 23andMe, and UK Biobank and eleven SNPs associated with cannabis use disorder (P-value < 5×10^−7^) from a GWAS meta-analysis of (17,068 cases and 357,219 controls of European descent) from Psychiatric Genomics Consortium Substance Use Disorders working group, Lundbeck Foundation Initiative for Integrative Psychiatric Research, and deCode. We associated these SNPs with the largest to date GWAS meta-analysis of POAG (16,677 cases and 199,580 controls).

**Results:** MR analysis suggested no evidence for a causal association of lifetime cannabis use and cannabis use disorder with POAG (odds ratio (OR) of outcome per doubling of the odds of the exposure (95% confidence interval): 1.04 (0.88; 1.23) for lifetime cannabis use and 0.97 (0.92; 1.03) for cannabis use disorder). Sensitivity analyses to address pleiotropy and weak instrument bias yielded similar estimates to the primary analysis.

**Conclusions:** Our results do not support a causal association between cannabis use and POAG.

## Introduction

Glaucoma is the leading cause of irreversible blindness worldwide, and is estimated to affect more than 100 million in 2040 [1]. Primary open-angle glaucoma (POAG), the most common subtype of glaucoma, is a slowly progressing optic neuropathy that can remain undetected for years, for which intraocular pressure (IOP) has been identified as the most significant modifiable risk factor [2]. In most of POAG cases the first line of therapy for lowering of the IOP, and thus slowing down POAG progression, is topical treatment with eye drops [3], which carry numerous risks for ocular side effects and usually require lifelong continuation [4].

Given the high incidence of glaucoma and the limitations of the current anti-glaucoma agents, research has focused during the last decades on the identification of novel treatment modalities, including the use of cannabis [5]. Cannabinoids have been found to exert a lowering effect on IOP when administered intravenously, orally, or by smoking [5]. However long-term and adequately sized clinical trials testing cannabinoids treatment in POAG are lacking. Moreover, in the existing observational studies assessing the association between cannabis smoking and IOP, and subsequently on the risk of POAG, it is challenging to isolate the individual effects of cannabinoids and tobacco, since they are usually consumed together [6].

One approach to strengthen the causal inference on the association of cannabis use and the risk of POAG is Mendelian randomization (MR), a form of instrumental variable analysis that uses genetic variants as instruments [7]. In the present study, we used MR to assess any potential causal association between cannabis use and the risk of POAG.

## Materials and Methods

### Study design

MR uses genetic variants as instrumental variables to assess causal associations between risk factors and diseases based on the random assignment of genetic variants in individuals at conception [7]. These genetic variants are usually single-nucleotide polymorphisms (SNPs) and since they are randomly allocated in individuals independently of other factors, MR studies can serve as naturally occurring randomized controlled trials [7]. Thus, MR association estimates are less prone to biases occurring from confounding and reverse causation than those derived from traditional observational studies. We performed a two-sample, summary-based MR and we utilized summary statistics from three genome-wide association studies (GWAS) of lifetime cannabis use [8], cannabis use disorder [9] and POAG [10]. Then by combining these estimates we calculated the causal association between cannabis use and cannabis use disorder with POAG. We followed the recommendations by STROBE-MR [11] and “Guidelines for performing Mendelian randomization investigations” [7]. The study protocol was pre-registered.

### Data sources

We retrieved summary data from the largest GWAS to date for lifetime cannabis use comprising 184,765 individuals of European descent, by the International Cannabis Consortium, 23andMe, and UK Biobank [8]. The exposure was defined as any self-reported use of cannabis during lifetime. GWAS analysis were adjusted for sex, age, ancestry, and genotype batch. Genotyping and imputation methods have been described elsewhere [8]. We also retrieved summary statistics for cannabis use disorder from a GWAS meta-analysis of 17,068 cases and 357,219 controls of European descent, derived from the Psychiatric Genomics Consortium Substance Use Disorders working group, Lundbeck Foundation Initiative for Integrative Psychiatric Research (iPSYCH), and deCODE (Supplementary Table 1) [9]. Cases from the Psychiatric

Genomics Consortium had the diagnosis of cannabis abuse or dependence according to the Diagnostic and Statistical Manual of Mental Disorders (DSM)-IV or DSM-III-R, from clinician ratings or semi-structured interviews. IPSYCH cases met the criteria for a diagnosis of cannabis abuse (F12.1) or cannabis dependence (F12.2) based on the ICD-10 criteria, while cases from the deCODE sample have been diagnosed with lifetime cannabis abuse or dependence according to DSM-IV or DSM-III-R, or with DSM-V cannabis use disorder. Genotyping, quality control and imputation methods have been described elsewhere [9]. SNP-POAG associations were taken from GWAS meta-analysis of 16,677 POAG cases and 199,580 controls of European ancestry [10] from 16 participating studies (Supplementary Table 1). POAG was defined according to ICD9/ICD10 criteria. GWAS adjusted for age, sex, and study-specific principal components [10]. Genotyping, quality control and imputation have been described in detail elsewhere [10].

### Selection of genetic variants as instrumental variables

We adopted two approaches in the selection of genetic variants as instrumental variables. In the primary analysis we selected only SNPs reaching genome-wide significance (P-value < 5*10^−8^ for lifetime cannabis use and P-value < 5*10^−7^ for cannabis use disorder) following clumping for linkage disequilibrium (LD) at r^2^ < 0.001 across a 10mb window. In our secondary, more liberal approach [12, 13], we selected SNPs independently associated with lifetime cannabis use and cannabis use disorder at a GWAS P-value < 5*10^−5^ after accounting for LD at r^2^ < 0.1, in order to increase pooled instrument strength and power of the analysis. Finally, we performed the MR-Steiger directionality test to identify the direction of causality between lifetime cannabis use and POAG and removed SNPs that were more strongly correlated with the outcome than the exposure [14]. We excluded SNPs with highly influential data points in the funnels plots of single SNP Wald ratio estimates and scatter plots of SNP–exposure associations versus SNP– outcome associations. Five SNPs associated with lifetime cannabis use and eleven SNPs associated with cannabis use disorder were selected in the primary analysis, while 267 and 157 SNPs associated with lifetime cannabis and cannabis use disorder, respectively, were selected in the secondary analysis.

### Statistical analysis

After data harmonization, where SNPs were filtered according to HapMap3 [15], excluded if they were strand-ambiguous and their effect sizes were aligned, we calculated Wald ratios by dividing the per-allele logarithm of odds ratio (logOR) for each selected SNP from the lifetime cannabis use and cannabis use disorder GWAS by the corresponding logOR from the same SNP in the POAG GWAS. Then, we estimated the effect of lifetime cannabis use and cannabis use disorder on the risk of POAG by pooling the Wald ratios with multiplicative random effects inverse-variance weighted (IVW) meta-analyses [12].

Univariable two-sample MR was performed using summary-level statistics from the largest available GWAS on lifetime cannabis use and POAG. The two-sample MR approach rests on 3 core assumptions: (1) the genetic instruments should be robustly associated with the exposure of interest (“relevance” assumption), (2) the genetic instruments are not associated with confounders of the exposure-outcome association (“exchangeability” assumption), and (3) the genetic instruments are associated with the outcome exclusively through their effect on the exposure of interest (“exclusion restriction” assumption) [16, 17]. The “relevance” assumption is satisfied by selecting SNPs, as instrumental variables, reaching the genome-wide significance (P-value < 5*10^−8^). Moreover, in order to quantify instrument strength, we calculated the proportion of variance of the exposure explained by the genetic instruments, as well as the F-statistic of our instruments [18]. Although, the “relevance” and “exclusion restriction” assumption cannot be proven, we performed sensitivity analyses to assess any possible violations of these assumptions. These can occur through horizontal pleiotropy, where the genetic variants affect the outcome via biological pathways other than the exposure under investigation. Thus, in the primary analysis, we utilized PhenoScanner [19] to investigate associations between the selected genetic instruments with traits that could potentially confound our analysis and in case that pleotropic pathways were discovered, multivariable MR was used to adjust for these effects [20]. Additionally, the associations of each selected SNP and its proxies (r^2^ > 0.8) with known risk factors for POAG were also checked. In the multivariable MR analyses the conditional F-statistic was used as a quantification of the strength of our genetic instruments [21]. Moreover, we assessed the heterogeneity among the selected genetic variants in the primary analysis through the Cochran Q heterogeneity test and I _GX_^2^ [17] in order to detect pleiotropy. MR Egger regression was performed in order to assess the presence of directional pleiotropy [17], as well as pleiotropy-robust methods [22] (penalized weighted median, IVW radial regression and MR-Pleiotropy Residual Sum and Outlier (MR-PRESSO). Since only five SNPs for lifetime cannabis use were selected in our primary analysis, the IVW radial regression and the MR-PRESSO were not performed [22]. In order to assess whether the IVW estimate was driven by a single SNP, leave-one-out analysis was also conducted.

After applying the liberal instrument selection strategy, we performed multiplicative random-effects IVW and pleiotropy-robust methods (penalized weighted median, IVW radial regression, MR-Pleiotropy Residual Sum and Outlier (MR-PRESSO)) [22]. The CAUSE MR analysis was additionally conducted as an additional method to improve statistical power and mitigate the risk of weak instrument bias [7, 23].

All MR estimates for the associations between our exposures and POAG were multiplied by log_e_2 (=0.693), representing the change in log odds of POAG per doubling in the prevalence of our exposures [24]. All analyses were performed with R version 4.2.1 [25] using the cause (1.2.0.335), MendelianRandomization (0.5.1), MVMR (0.3), MR-PRESSO (1.0), TwoSampleMR (0.5.6), GenomicSEM, LCV and lhcMR packages.

## Results

In our primary analysis the selected 5 SNPs explained 0.09% of the variance in the lifetime cannabis use and the F-statistics for all SNPs were ≥ 30.7. The selected 11 SNPs for cannabis use disorder explained 0.08**%** of the phenotypic variance and had an F-statistic of ≥ 25.5. We found no evidence for an effect of the genetically predicted lifetime cannabis use on the POAG risk using the IVW method (OR=1.04 per doubling odds of exposure; 95%CI = 0.88 to 1.23; P-value = 0.67). The estimate from the penalized weighted median analysis was consistent with the estimate from the IVW analysis (Figure 1). Similarly, estimates from the IVW analysis, as well as the pleiotropy-robust methods, did not support an association between genetically predicted cannabis use disorder and POAG (OR=0.97 per doubling odds of exposure; 95% CI:0.92 to 1.03; P-value = 0.27) (Figure 2).

**Figure 1.**
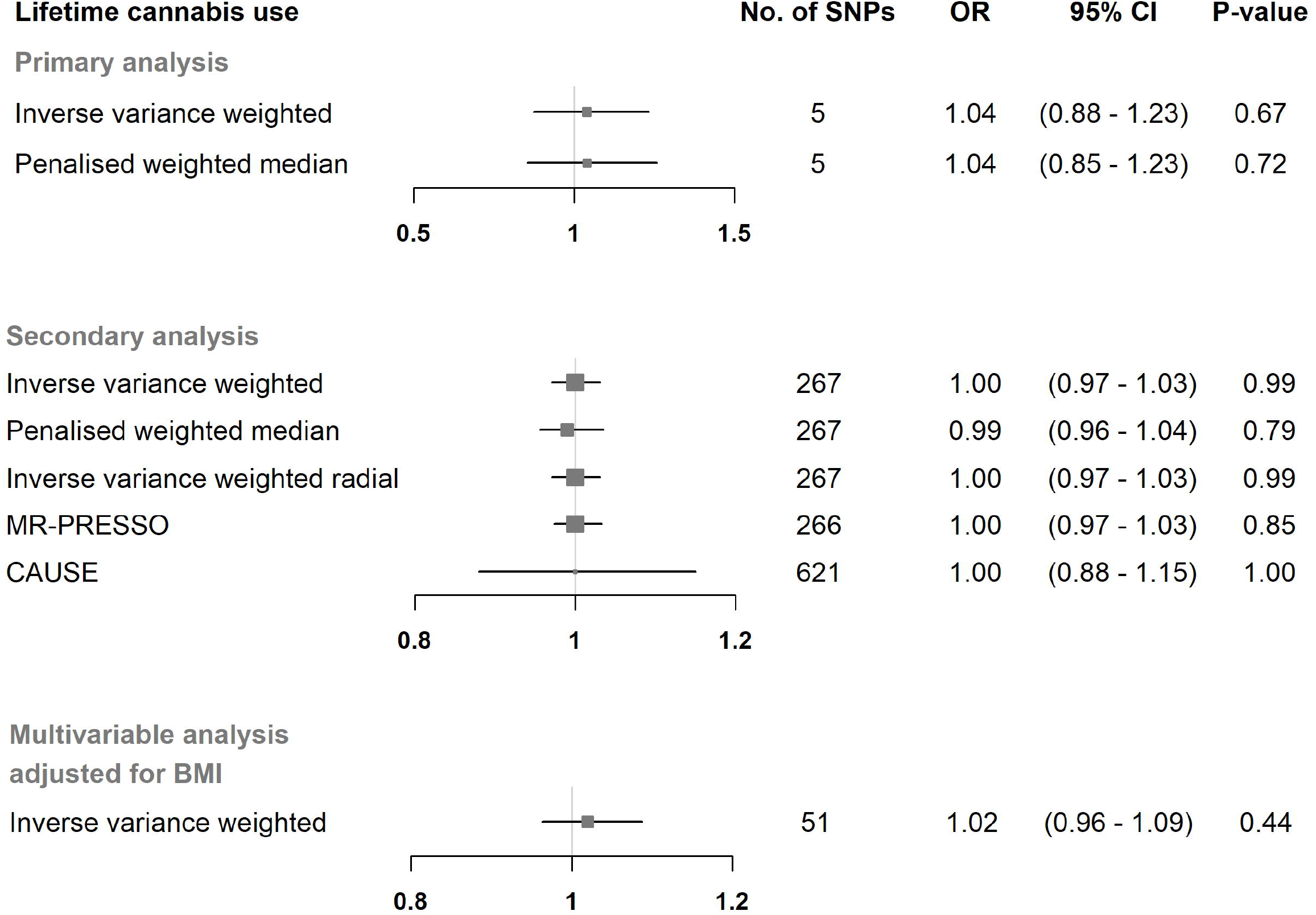
Mendelian randomization estimates for the effect of lifetime cannabis use on primary open-angle glaucoma. Estimates are reported as changes in odds of primary open-angle glaucoma per doubling in the prevalence of lifetime cannabis use. SNP, single nucleotide polymorphism; CI, confidence interval; MR-PRESSO, Mendelian randomization pleiotropy residual sum and outlier; CAUSE, causal analysis using summary effect estimates; BMI, body mass index

**Figure 2.**
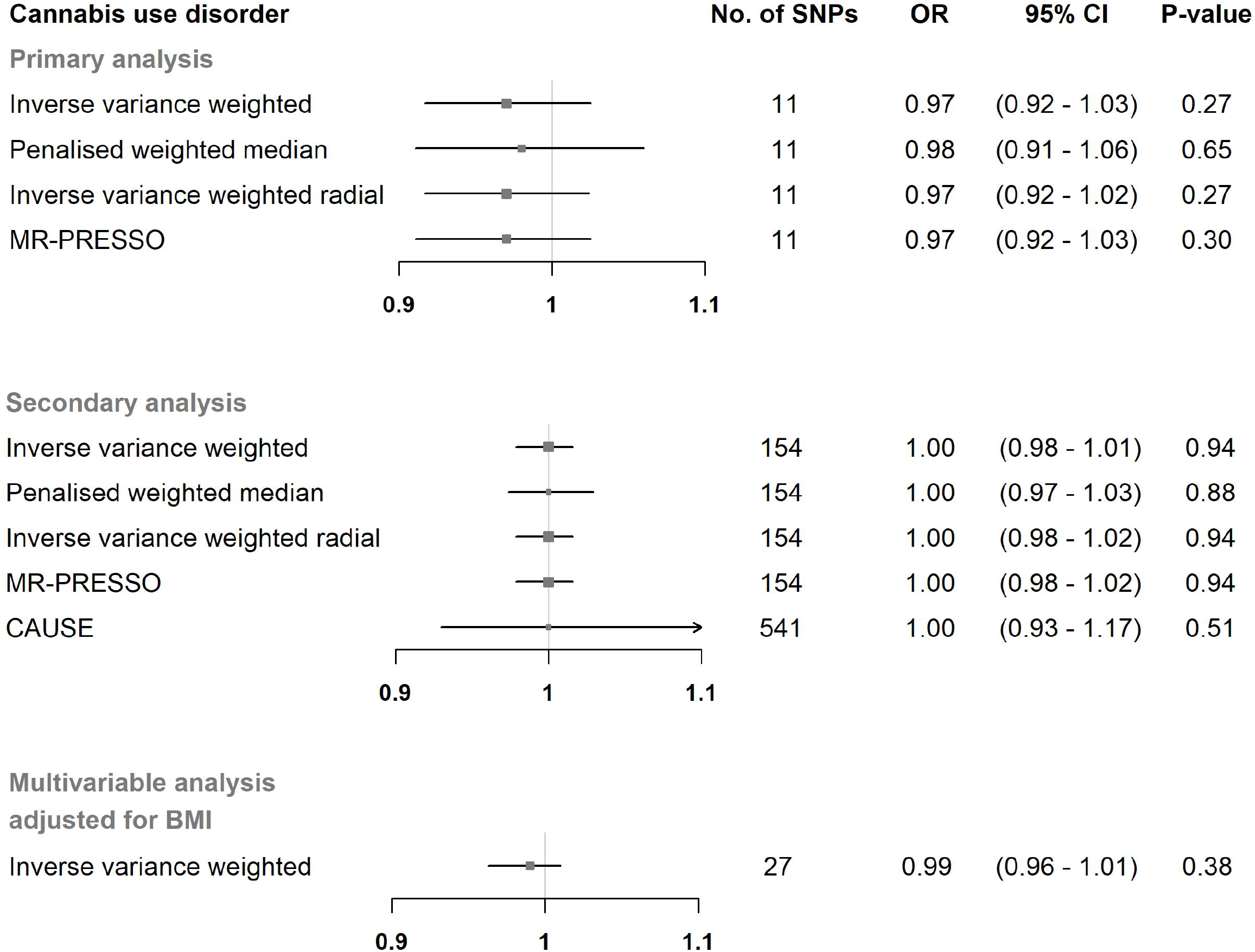
Mendelian randomization estimates for the effect of cannabis use disorder on primary open-angle glaucoma. Estimates are reported as changes in odds of primary open-angle glaucoma per doubling in the prevalence of cannabis use disorder. SNP, single nucleotide polymorphism; CI, confidence interval; MR-PRESSO, Mendelian randomization pleiotropy residual sum and outlier; CAUSE, causal analysis using summary effect estimates; BMI, body mass index

One of our instrumental SNPs for lifetime cannabis use was associated with previously reported obesity-related phenotypes (Supplementary Table 3). Several studies have found an association between BMI and POAG [26, 27], so this SNP might have been associated with the POAG through pathways other than our exposure of interest and, thus, we performed multivariable IVW adjusting for body mass index (BMI). In the multivariable IVW analysis adjusted for BMI we found no evidence of horizontal pleiotropy introduced to the univariable estimates from lifetime cannabis use and cannabis use disorder (Figure 1 and 2). The conditional F-statistics for lifetime cannabis use and cannabis use disorder were 12.5 and 11.6, respectively.

In our primary analysis there was no evidence of heterogeneity among Wald ratios for lifetime cannabis use and cannabis use disorder with POAG (Supplementary Table 4). The intercepts from the MR-Egger analyses did not deviate from zero, thus, no directional pleiotropy was present (Supplementary Table 4). The leave-one-SNP-out analyses identified no SNPs with high influence on the IVW estimates for our exposures (Supplementary Table 5).

In our secondary analysis using a liberal threshold, the selected 267 SNPs explained 2.99% of the variance in the lifetime cannabis use and the F-statistics for all SNPs were ≥ 16.4. The selected 157 SNPs for cannabis use disorder explained 0.82% of the phenotypic variance and had an F-statistic of ≥ 16.4. MR estimates from the IVW analyses, as well as from the pleiotropy-robust models showed no association between our exposures and POAG (Figure 1 and 2). The MR-PRESSO global test provided evidence for one outlier SNP (p-value = 0.005) in the analysis with lifetime cannabis use as an exposure, which was removed (rs8140423) from the calculation of the final MR-PRESSO estimate. MR-PRESSO distortion test showed that the outlier corrected estimate did not differ significantly from the non-corrected estimate (p-value = 0.93).

The CAUSE models included more instrumental SNPs in order to increase statistical power and did not reveal a causal effect of lifetime cannabis and cannabis use disorder on POAG (Figure 1 and 2).

## Discussion

In this two-sample MR we leveraged genetic data of lifetime cannabis use and cannabis use disorder from more than 180,000 and 370,000 individuals, respectively, and of 16,000 POAG cases, to assess the association of cannabis user with the risk of POAG. We found no evidence to support the hypothesis that cannabis use affects the development of POAG.

During the last decades, several lines of evidence have been put forward to elucidate the effect of cannabis on POAG. It has been postulated that cannabis consumption may have a protective effect on the risk of POAG, by a salutary effect on IOP [5], without knowing the exact pathogenetic mechanism of this phenomenon. It has been hypothesized that IOP decrease results from the activation of the cannabinoid-related receptors in the ciliary body of the eye, by Δ^9^-tetrahydrocannabinol, the psychoactive constituent of cannabis [28]. As a result, the ciliary body reduces the production of aqueous humor and IOP decreases. An alternative hypothesis supports that the IOP lowering effect of cannabis is mediated through the decrease in blood pressure [29]. However, this hypothesized mechanism could potentially increase the development of POAG since it lowers ocular perfusion pressure and, thus, compromises adequate perfusion of optic nerve head [30]. Moreover, when cannabis is smoked, several toxic and carcinogenic compounds are inhaled [31]. The systemic absorption of these compounds may occur in higher concentrations than in tobacco smoking, mainly because of the way that cannabis is smoked. Usually, filters are lacking in cannabis cigarettes and a longer and deeper inhalation is required [32]. As a result, cannabis smoking may yield similar negative effects as tobacco smoking in POAG [33].

Although, several interventional studies have investigated the effects of cannabis in IOP, evidence on the effect of cannabis in POAG are scarce and usually limited by small sample sizes. In a recent prospective study of the UK Biobank cohort [34], lifetime cannabis use has not been associated with POAG in multivariable analysis adjusted for tobacco smoking and other confounders. Similar to our MR estimates, the OR of POAG for cannabis use versus never-cannabis use was 1.03 (95% CI: 0.91–1.17). On the contrary, in the same study, it has been found that participants who have used cannabis 11 to 100 times in their lifetime had lower mean IOP compared to those who have never consumed cannabis, without, however, having adjusted for any potential confounders. The existing interventional studies on the association between cannabis use and IOP [35] have several limitations including, small sample sizes and duration of studies, inclusion of patient with various types of glaucoma, which limits the generalization of the result, and lack of specification of the time that IOP was measured.

The key strength of this study was the utilization of the largest to date GWAS meta-analysis of POAG, which increased the statistical precision of our estimates. Moreover, our MR estimates have been shown to be robust to model violations in sensitivity analyses. The study has also several limitations. First, because of the binary nature of our exposure we were not able to assess any dose-dependent changes in POAG. A more detailed description of our exposure was not available so we were also able to assess neither the chemical composition of cannabis consumed nor its route of administration. Second, we did not investigate the association of cannabis use on other types of glaucoma (e.g., low tension glaucoma).

In conclusion, our data provided evidence for a lack of association of genetic liability to lifetime cannabis use and cannabis use disorder with POAG. Triangulation of evidence from different types of research studies, with different key sources of bias, is warranted to confirm our results.

## Data Availability

The summary statistics for the lifetime cannabis use cannabis GWAS are available at https://www.ru.nl/bsi/research/group-pages/substance-use-addiction-food-saf/vm-saf/genetics/international-cannabis-consortium-icc/ (access date: 2022/10/17). The cannabis use disorder data are available at https://ipsych.dk/en/research/downloads/data-download-agreement-ipsych-secondary-phenotypes-cannabis-2019/ (access date: 2022/10/17). The primary open-angle glaucoma summary data are available at https://www.ebi.ac.uk/gwas/publications/33627673 (access date: 2020/07/20) and the summary data from the body mass index GWAS is available from https://portals.broadinstitute.org/collaboration/giant/index.php/GIANT_consortium_data_files#2018_GIANT_and_UK_BioBank_Meta-analysis(access date: 2022/10/17).

