## Supplementary tables and figures for "Cannabis use and the risk of primary open-angle glaucoma: a Mendelian randomization study"

Authors et al.

Supplementary Figures and Tables

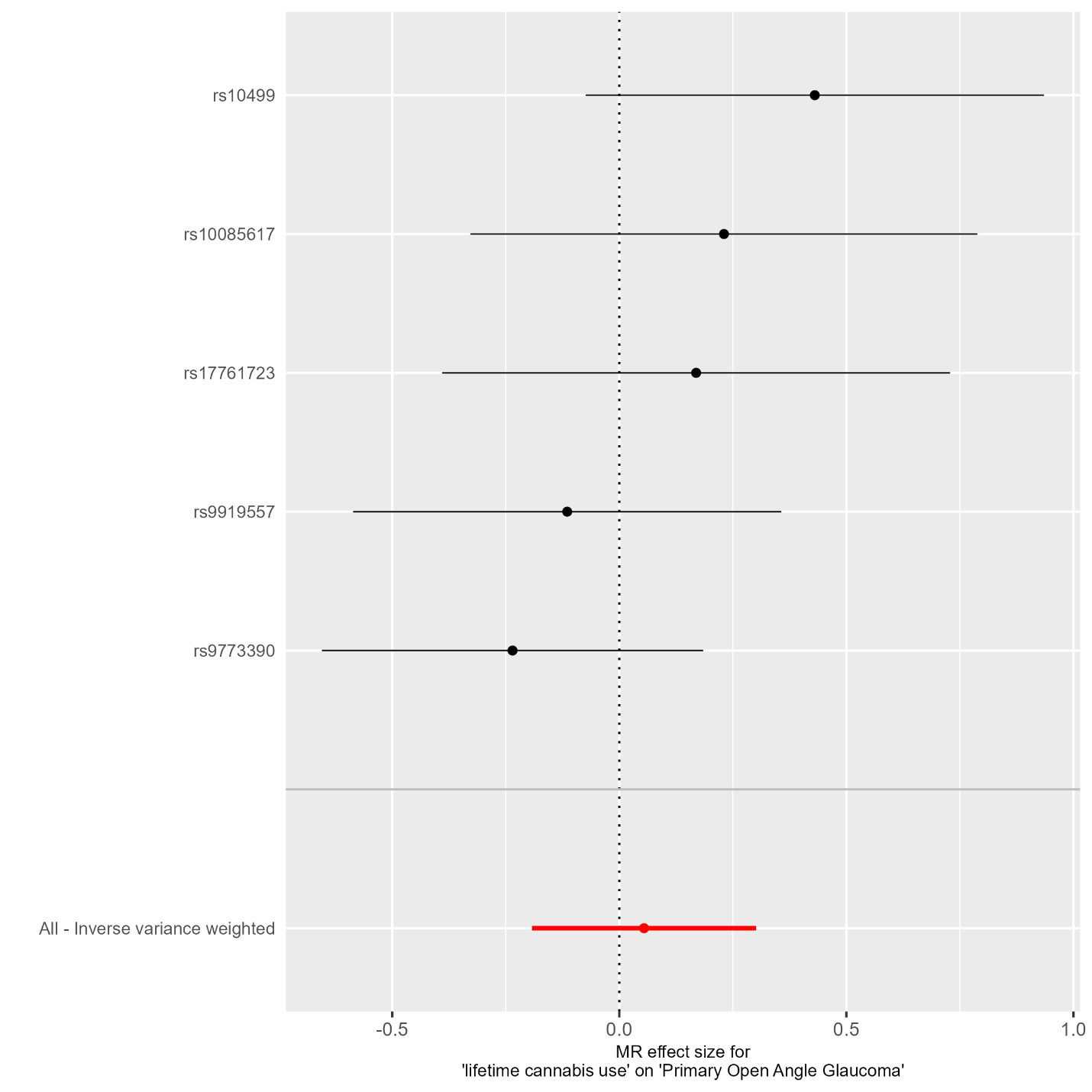
Supplementary Figure 1	Funnel plot of single SNP Wald ratio estimates for the effect of lifetime cannabis use on primary open-angle glaucoma

SNP: single-nucleotide polymorphism

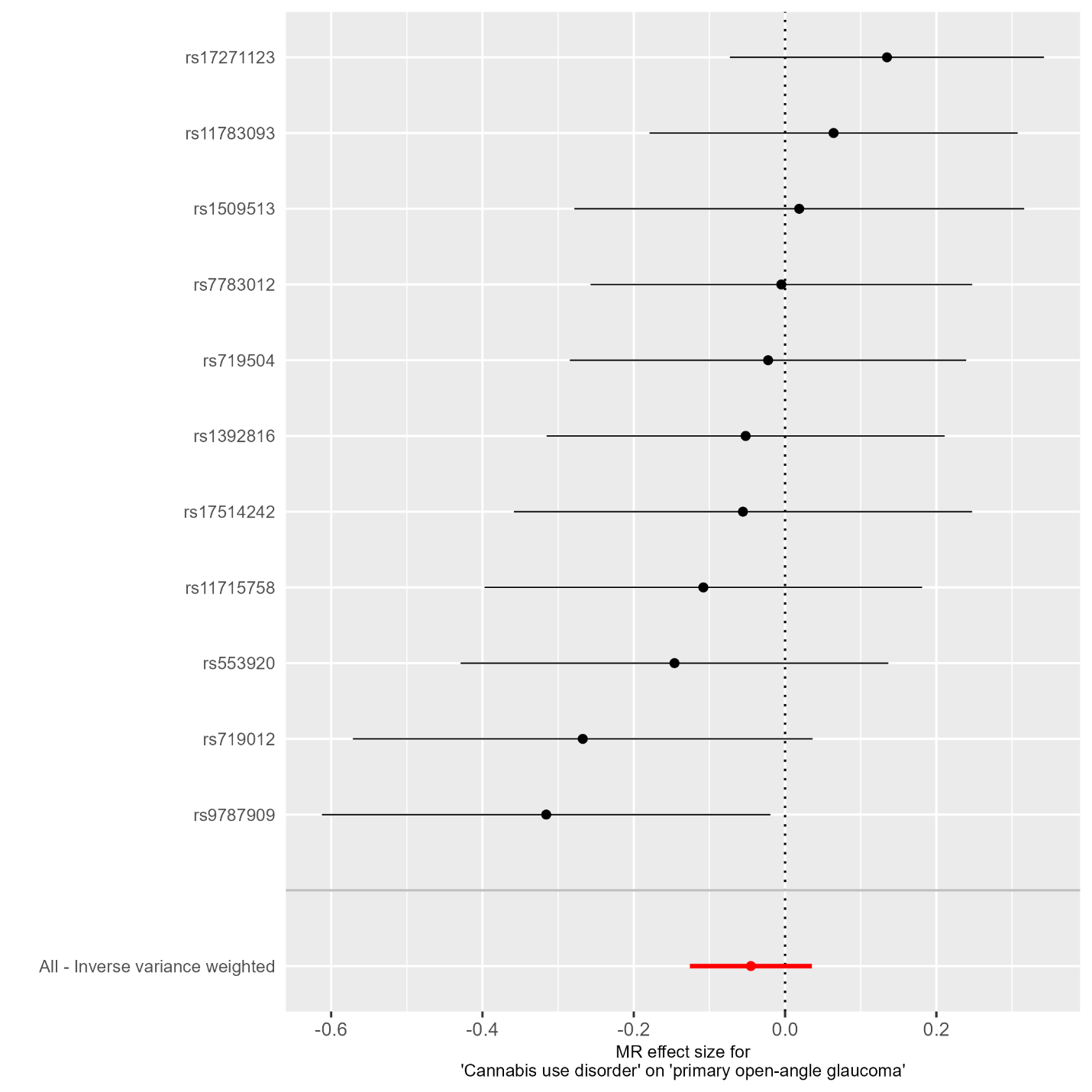
Supplementary Figure 2	Funnel plot of single SNP Wald ratio estimates for the effect of cannabis use disorder on primary open-angle glaucoma

SNP, single-nucleotide polymorphism

Supplementary Figure 3	Scatter plot of SNP-primary open-angle glaucoma associations vs SNP-lifetime cannabis use associations

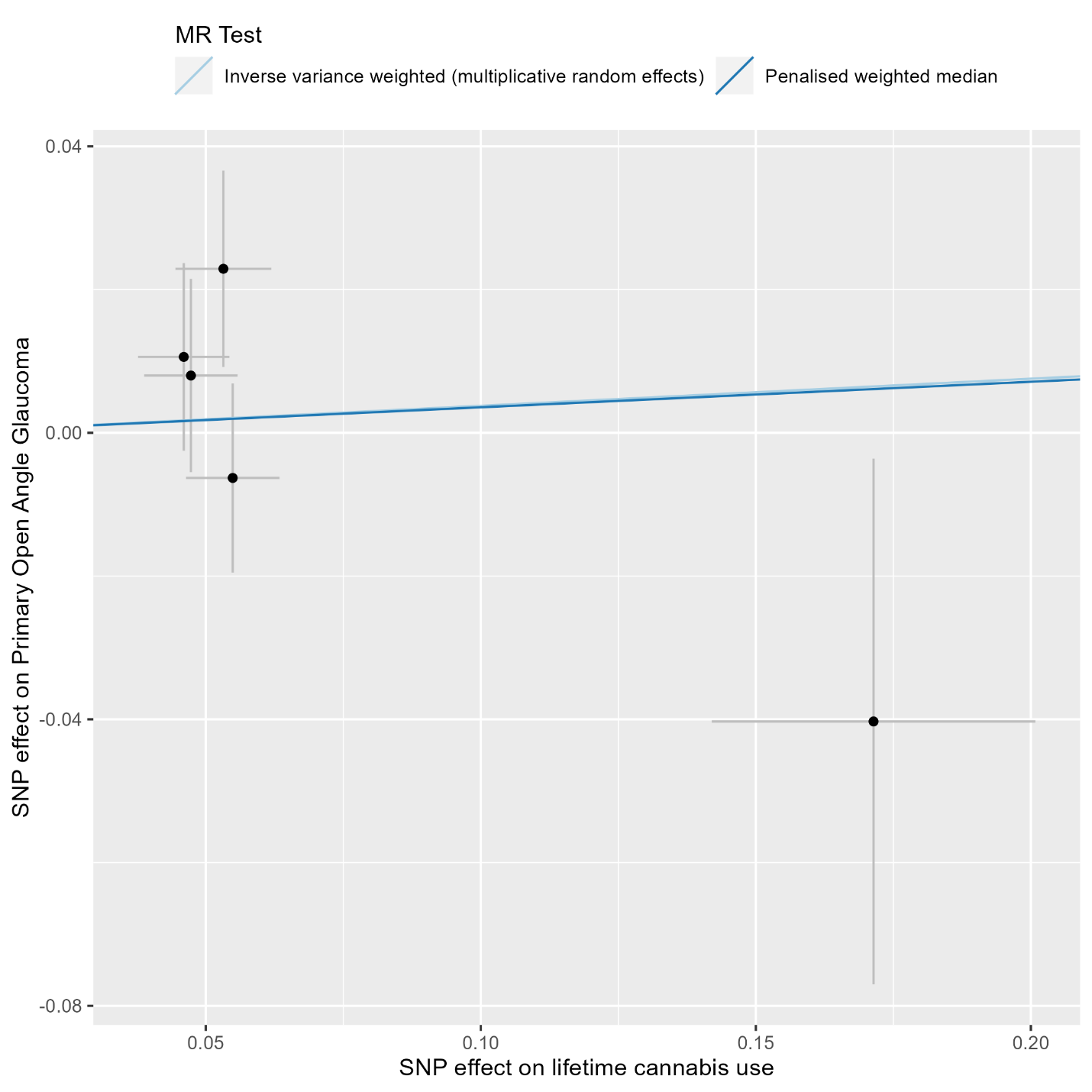

SNP: single-nucleotide polymorphism

Supplementary Figure 4	Scatter plot of SNP-primary open-angle glaucoma associations vs SNP-cannabis use disorder associations

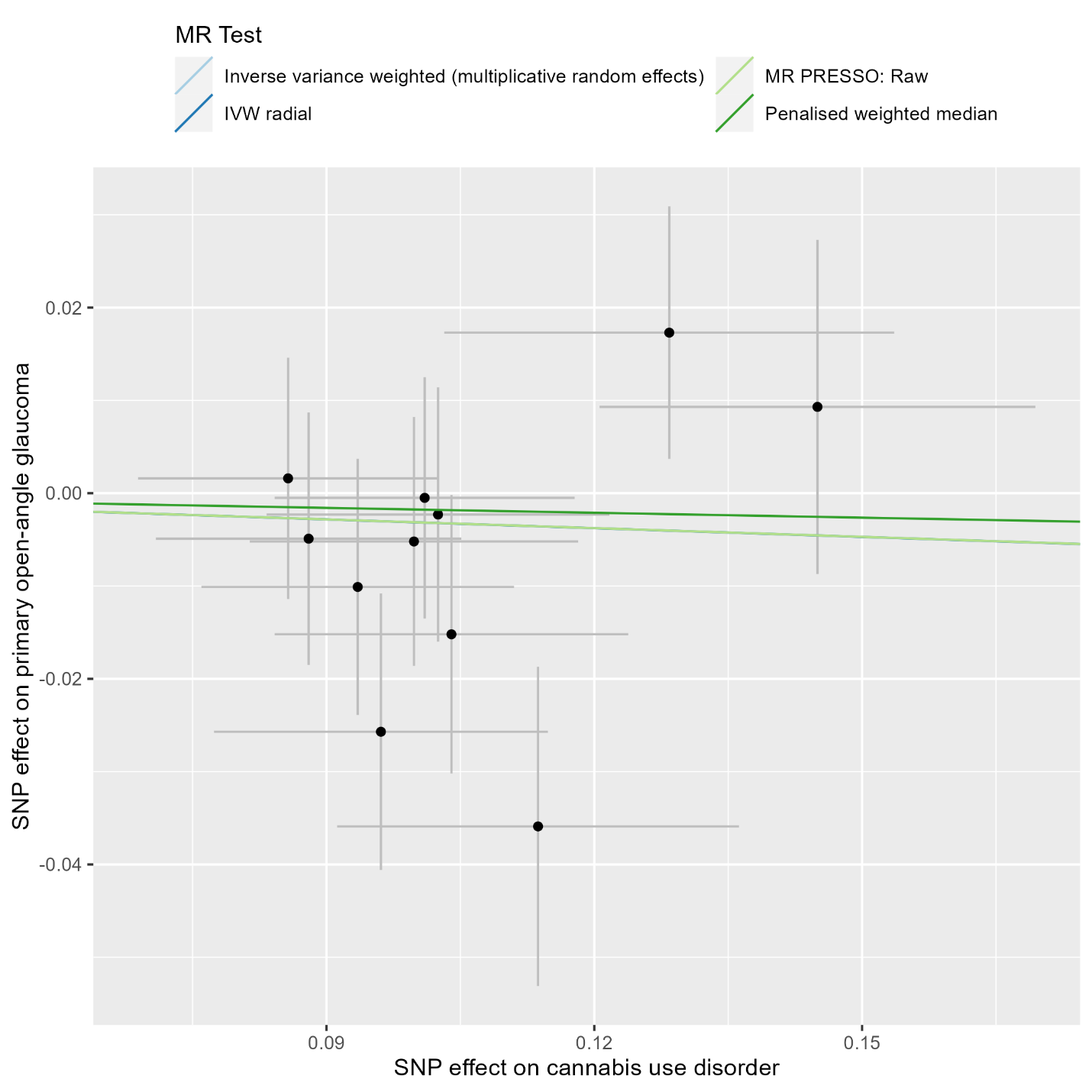

SNP: single-nucleotide polymorphism

Supplementary Table 1	Phenotypic descriptive statistics of studies included in the exposure, confounder/outcome risk factor, and outcome genome-wide association studies

| GWAS / Study | Phenotype | | | |
| --- | --- | --- | --- | --- |
| (Gharahkhani et al., 2021) | Primary open angle glaucoma | | | |
|  | N cases | N controls | Age (mean ± SD) cases | Age (mean ± SD) controls |
| NEIGHBOR/MEEI | 2606 | 2606 | 65.7 ± 13.4 | 68.3 ± 11.4 |
| EPIC-Norfolk Eye Study | 664 | 5630 | 61.5 ± 7.9 | 55.6 ± 7.9 |
| ANZRAG | 3071 | 6750 | NA | NA |
| UKBB POAG ICD10 code | 1448 | 22107 | 62.67 ± 5.6 | 56.36 ± 7.9 |
| Kaiser Permanente GERA Cohort | 3819 | 47961 | 79.6 ± 9.4 | 69.7 ± 13.0 |
| KCL | 576 | 287 | 70.5±13.5 | 80.3±4.3 |
| BMES | 107 | 600 | 68.9 ± 7.9 | 63.8 ± 8.3 |
| Southampton | 941 | 1557 | >40 | >40 |
| GHS | 47 | 2731 | 62.21 ±10.63 | 55.66 ±10.86 |
| HPFS illumina | 116 | 527 | 58.3 ± 8.4 | 51.5 ± 8.0 |
| HPFS Affy | 34 | 1297 | 58.7 ± 8.0 | 52.8 ± 7.9 |
| NHS illumina | 266 | 1692 | 55.8 ± 6.4 | 53.2 ± 6.5 |
| NHS Affy | 46 | 1992 | 55.7 ± 6.0 | 53.0 ± 6.7 |
| ERF | 110 | 1999 | NA | NA |
| Rotterdam Study I | 198 | 1282 | NA | NA |
| FINNGEN | 1824 | 93036 | 65.6 ± 11.9 | 57.8 ± 16.1 |
| Geisinger 60K | 664 | 5904 | >18 | >18 |
| Geisinger 30K | 140 | 1622 | >18 | >18 |
| (Pasman et al., 2018) | Lifetime cannabis use | | | |
|  | N | Female (%) | Age (Mean) | Cannabis users (%) |
| ALSPAC | 2,976 | 55.7 | 17.75 | 41.6 |
| BLTS | 721 | 57.1 | 26.2 | 59.5 |
| CADD | 853 | 30.2 | 24.9 | 79.3 |
| EGCUT1 | 2,765 | 54.5 | 33.8 | 1.3 |
| EGCUT2 | 970 | 51.2 | 31.1 | 4.8 |
| FinnTwin | 1,029 | 51.7 | 22.8 | 27.4 |
| HUVH | 981 | 29.8 | 35.6 | 20.1 |
| MCTFR | 6,241 | 53.5 | 37.2 | 59.3 |
| NTR | 4,653 | 66.2 | 36.9 | 26.9 |
| QIMR | 6,778 | 53.8 | 45.2 | 51.4 |
| TRAILS | 1,226 | 46.9 | 19.0 | 50.5 |
| Utrecht | 1,173 | 53.5 | 20.6 | 54.1 |
| Yale Penn EA | 1,964 | 40.2 | 38.2 | 91.6 |
| Radar | 338 | 44.4 | 19.54 | 58.8 |
| SYS | 551 | 56 | 49.52 | 52.45 |
| TwinsUK | 2,070 | 92.6 | 57.7 | 11.7 |
| Yale Penn AA | 2,660 | 46.4 | 41.6 | 81.8 |
| 23andme | 22,683 | 55.3 | 54 | 43.2 |
| UKB | 126,785 | 56.3 | 55 | 22.3 |
| (Johnson et al., 2020) | Cannabis use disorder | | | |
|  | Cases | Controls |  |  |
| CATS | 958 | 453 |  |  |
| CADD | 397 | 699 |  |  |
| CHDS | 201 | 420 |  |  |
| FSCD | 226 | 314 |  |  |
| COGEND Nico | 306 | 607 |  |  |
| COGEND SAGE | 228 | 830 |  |  |
| GEDI-GSMS | 81 | 491 |  |  |
| BLTS | 170 | 1216 |  |  |
| MCTFR | 449 | 1625 |  |  |
| Yale Penn 1 | 916 | 833 |  |  |
| Yale Penn 2 | 557 | 497 |  |  |
| bigCOGA | 2206 | 5053 |  |  |
| CEDAR | 64 | 148 |  |  |
| OZ-ALC | 593 | 4893 |  |  |
| VTSABD | 99 | 734 |  |  |
| IASPSAD | 104 | 613 |  |  |
| Add health | 722 | 4071 |  |  |
| iPSYCH | 2758 | 53326 |  |  |
| deCODE | 6033 | 280396 |  |  |
| (Pulit et al., 2019) | Body mass index | | | |
|  | N | BMI (Mean ± SD) | Female (%) |  |
| UKB + GIANT | 806,834 | 27.4 (4.8) | 54 |  |

|  |  |  |  | Estimates for exposure | | | | Estimates for primary open angle glaucoma | | |
| --- | --- | --- | --- | --- | --- | --- | --- | --- | --- | --- |
| SNP | EA | OA | EAF | BETA | SE | P | F | BETA | SE | P |
| Lifetime cannabis use | | | | | | | | | | |
| rs10085617 | A | T | 0.416 | 0.046 | 0.008 | 2.9e-08 | 30.7 | 0.011 | 0.013 | 0.209 |
| rs10499 | A | G | 0.651 | 0.053 | 0.009 | 1.1e-09 | 37.4 | 0.023 | 0.014 | 0.047 |
| rs17761723 | T | C | 0.346 | 0.047 | 0.009 | 3.2e-08 | 31.0 | 0.008 | 0.014 | 0.277 |
| rs9773390 | T | C | 0.933 | -0.171 | 0.029 | 5.7e-09 | 34.0 | 0.040 | 0.037 | 0.136 |
| rs9919557 | T | C | 0.614 | -0.055 | 0.009 | 9.9e-11 | 41.7 | 0.006 | 0.013 | 0.317 |
| Cannabis use disorder | | | | | | | | | | |
| rs11715758 | A | G | 0.622 | -0.094 | 0.018 | 8.9e-08 | 28.5 | 0.010 | 0.014 | 0.232 |
| rs11783093 | T | C | 0.846 | -0.145 | 0.024 | 2.7e-09 | 35.3 | -0.009 | 0.018 | 0.303 |
| rs1392816 | T | C | 0.624 | -0.100 | 0.018 | 6.1e-08 | 29.4 | 0.005 | 0.013 | 0.349 |
| rs1509513 | A | G | 0.546 | 0.086 | 0.017 | 3.2e-07 | 26.0 | 0.002 | 0.013 | 0.451 |
| rs17271123 | T | G | 0.587 | 0.128 | 0.025 | 3.5e-07 | 26.0 | 0.017 | 0.014 | 0.102 |
| rs17514242 | C | G | 0.649 | -0.088 | 0.017 | 2.6e-07 | 26.5 | 0.005 | 0.014 | 0.359 |
| rs553920 | T | C | 0.770 | 0.104 | 0.020 | 1.6e-07 | 27.6 | -0.015 | 0.015 | 0.155 |
| rs719012 | T | C | 0.264 | 0.096 | 0.019 | 2.9e-07 | 26.4 | -0.026 | 0.015 | 0.042 |
| rs719504 | A | G | 0.650 | 0.102 | 0.019 | 9.0e-08 | 28.5 | -0.002 | 0.014 | 0.433 |
| rs7783012 | A | G | 0.476 | 0.101 | 0.017 | 1.8e-09 | 36.1 | 0.000 | 0.013 | 0.485 |
| rs9787909 | A | C | 0.829 | 0.114 | 0.022 | 4.5e-07 | 25.5 | -0.036 | 0.017 | 0.018 |
| EA, effect allele. OA, other allele. EAF, effect allele frequency. SE, standard error. | | | | | | | | | | |

Supplementary Table 2	Associations of single nucleotide polymorphisms for lifetime cannabis use and cannabis use disorder

Supplementary Table 3	Association (P<5x10^-8^) of the single nucleotide polymorphisms used as instruments with confounders or outcome risk factors in PhenoScanner (accessed on 2022/11/20 using the phenoscanner function of the R MendelianRandomization package)

| SNP | Phenotypes | PMID |
| --- | --- | --- |
| Lifetime cannabis use | |  |
| rs10499 | Mean corpuscular volume | 27863252 |
| rs10499 | Red blood cell count | 27863252 |
| rs10499 | Hip circumference | 25673412 |
| rs10499 | Waist circumference | 25673412 |
| rs10499 | Crohns disease | 26192919 |
| Cannabis use disorder | | |
| rs11715758 | High light scatter percentage of red cells | 27863252 |
| rs11715758 | High light scatter reticulocyte count | 27863252 |
| rs11715758 | Immature fraction of reticulocytes | 27863252 |
| rs11715758 | Reticulocyte count | 27863252 |
| rs11715758 | Reticulocyte fraction of red cells | 27863252 |
| rs7783012 | Years of educational attainment | 27225129 |
| rs7783012 | Age first birth | 27798627 |

PMID, PubMed ID. Body mass index was considered as a relevant confounder or risk factor for primary open angle glaucoma.

Supplementary Table 4	Heterogeneity of Wald ratios and MR-Egger test for directional pleiotropy

| Lifetime cannabis use | Heterogeneity | | |  |
| --- | --- | --- | --- | --- |
|  | Q | Degrees of Freedom | P | I_GX_^2^ |
| Lifetime cannabis use | 2.8 | 4 | 0.592 | 0.1 |
| Cannabis use disorder | 9.9 | 10 | 0.45 | 0.036 |
|  | MR-Egger test for directional pleiotropy | | |  |
|  | Intercept | Standard error | P |  |
| Lifetime cannabis use | 2.927e-02 | 0.018 | 0.197 |  |
| Cannabis use disorder | -3.637e-02 | 0.029 | 0.239 |  |

Supplementary Table 5	Inverse variance weighted estimates in leave-one-out analysis in primary analysis

| SNP excluded | SNP | OR | (95% CI) | P value |
| --- | --- | --- | --- | --- |
| Lifetime cannabis use | rs10085617 | 1.02 | (0.83;1.25) | 0.885 |
|  | rs10499 | 0.98 | (0.84;1.14) | 0.757 |
|  | rs17761723 | 1.02 | (0.83;1.26) | 0.830 |
|  | rs9773390 | 1.12 | (0.95;1.32) | 0.169 |
|  | rs9919557 | 1.07 | (0.87;1.32) | 0.507 |
| Cannabis use disorder | rs11715758 | 0.97 | (0.92;1.03) | 0.369 |
|  | rs11783093 | 0.96 | (0.9;1.02) | 0.179 |
|  | rs1392816 | 0.97 | (0.91;1.03) | 0.326 |
|  | rs1509513 | 0.97 | (0.91;1.03) | 0.257 |
|  | rs17271123 | 0.95 | (0.9;1) | 0.042 |
|  | rs17514242 | 0.97 | (0.91;1.03) | 0.321 |
|  | rs553920 | 0.98 | (0.92;1.03) | 0.408 |
|  | rs719012 | 0.98 | (0.93;1.03) | 0.472 |
|  | rs719504 | 0.97 | (0.91;1.03) | 0.293 |
|  | rs7783012 | 0.97 | (0.91;1.03) | 0.272 |
|  | rs9787909 | 0.98 | (0.94;1.03) | 0.514 |
